# Structural variant calling and clinical interpretation in 6224 unsolved rare disease exomes

**DOI:** 10.1101/2023.10.28.23297720

**Authors:** German Demidov, Steven Laurie, Annalaura Torella, Giulio Piluso, Marcello Scala, Manuela Morleo, Vincenzo Nigro, Holm Graessner, Siddharth Banka, Solve-RD consortium, Katja Lohmann, Stephan Ossowski

## Abstract

Structural variants (SVs), including large deletions, duplications, inversions, translocations, and complex SVs have the potential to disrupt gene function resulting in rare disease. Nevertheless, current pipelines and clinical decision support systems for exome sequencing (ES) tend to focus on small alterations such as single nucleotide variants (SNVs) and insertions-deletions shorter than 50 base pairs (indels). Additionally, detection and interpretation of large copy-number variants (CNVs) are frequently performed. However, detection of other types of SVs in ES data is hampered by the difficulty of identifying breakpoints in off-target (intergenic or intronic) regions, which makes robust identification of SVs challenging. In this paper, we demonstrate the utility of SV calling in ES resulting in a diagnostic yield of 0.4% (23 out of 5,825 probands) for a large cohort of unsolved patients collected by the Solve-RD consortium. Remarkably, 8 out of 23 pathogenic SV were not found by comprehensive read-depth-based CNV analysis, resulting in a 0.13% increased diagnostic value.

## Introduction

Next-generation sequencing (NGS) is a method widely used for the detection of single-nucleotide variants (SNVs) and short indels (Deciphering Developmental Disorders Study, 2015) in the clinical diagnosis of rare genetic diseases. Detection of larger or complex variants with short read NGS is challenging as the reads with a length of 100-150bp cannot span across distantly located breakpoints. Nonetheless, methods for the detection of structural variants (SVs), including copy-number variants (CNVs) as well as copy-number neutral variants (such as inversions and balanced translocations) have been developed for genome sequencing (GS). Three main signals are used to detect SVs from genome data: 1) paired-end (PE) orientation and abnormal insert size (distance between read one and read two in a pair), 2) the presence of split and soft-clipped (SC) reads at the breakpoints of SVs, and 3) abnormal read depths (RD) in CNVs (Tattini et al., 2015).

Exome sequencing (ES) is a cost-effective alternative to GS. ES protocols include exon-enrichment by probe hybridization, followed by high-depth sequencing of the enriched exonic regions (typically 100x coverage for clinical exome sequencing). Thus, ES allows reliable detection of small coding and near-splice-site variants within these targeted regions, which are causative for the largest fraction of genetic diseases upon current knowledge. However, based on the enrichment strategy, ES almost completely lacks coverage of deep-intronic and intergenic variants, making the detection of variants, including SV breakpoints, in these genomic regions essentially impossible. Since it is much more likely for breakpoints of SVs to occur in the >98% non-exonic regions of the human genome, SVs are exceedingly hard to detect with ES data. Hence, usage of PE and SC signals is restricted to breakpoint-detection in regions covered by ES reads (Sadedin et al., 2018) and thus, SV detection in ES data is mainly limited to detection of CNVs (deletions and duplications) using the normalized RD signature. It is worth noting that the RD signal in ES suffers from many biases such as GC-content correlated coverage bias and does not allow a robust detection of short coding CNVs affecting only one to several exons, while short CNVs displaying PE or SC signals in addition to changes in RD are more reliably detectable. Therefore, a frequently used approach to SV detection in patients with negative ES results is to perform additional NGS analyses, such as short or long read GS, which increases both cost and time to diagnosis.

Despite the above-mentioned issues with SVs in ES data, for approximately 2-4% of SVs (depending on the exome kit’s target region size) we would expect the breakpoints to occur close enough to a targeted region, to be detected. In this paper, we evaluate the increase in diagnostic yield by PE- and SC-based SV calling in a large dataset of more than six thousand individuals with rare diseases (Solve-RD data freezes 1 and 2 (Zurek et al., 2021)) who remained undiagnosed after standard analysis of ES. Despite the large data heterogeneity, we show a valuable, albeit small increase in the diagnostic yield following SV calling, quality filtering, functional annotation, and clinical interpretation. We demonstrate the benefit of SV calling as part of ES reanalysis for patients with no previously identified molecular cause, underlining the value of applying SV calling algorithms in ES (re-)analysis projects.

## Materials and Methods

The recruitment of individuals investigated by the Solve-RD consortium has been described in detail in (Zurek et al., 2021). The part of the Solve-RD cohort used in this study includes 9,351 exome samples from 9,314 individuals (Solve-RD data freezes 1 and 2), including 6,224 affected individuals from 5,825 families and 3,090 unaffected relatives, usually parents. Samples were collected from multiple centers across Europe within four European Reference Networks (ERNs). Our cohort includes 1) 1,892 (30.4%) affected individuals from 1,821 families from ERN ITHACA (“Intellectual disability, TeleHealth And Congenital Anomalies”, including 26 affected individuals (0.4%) from 21 families from Undiagnosed Rare Diseases Program (Spain)), 2) 2,343 (37.6%) patients from 2,200 families from ERN RND (Rare Neurological Diseases), 3) 1,632 (26.2%) patients from 1,499 families from EURO-NMD (ERN for NeuroMuscular Diseases) and 4) 357 (5.7%) patients from 340 families from ERN-GENTURIS (GENetic TUmour RIsk Syndromes) (see Laurie et al, 2023, submitted, for more details). Patients were submitted by 44 clinical research groups from these 4 ERNs, who used 28 different exome enrichment kits (various vendors and kit versions). Human phenotype ontology (HPO) terms, family relationship information, and ES data for all patients were submitted to GPAP (Laurie et al., 2022) by the corresponding clinicians and submitters.

Reads were aligned to the GRCh37 hs37d5 human reference genome using BWA-MEM 0.7.8. Manta SV caller (Chen et al., 2016) was used to detect SVs using default parameters and exome flag on. The population allele frequency (AF) of a detected breakpoint was estimated across the complete dataset using the number of breakpoints detected within +/-20 base pairs to the focal breakpoint. This allowed removal of frequently observed events (polymorphisms) and artifacts. We kept only SVs with a breakpoint frequency of less or equal to 20 out of 9,351 exome datasets from affected and unaffected individuals. This AF threshold of °0.21% was selected empirically, considering the largest family sizes and previously reported highest frequencies of variants involved in recessive disorders. Moreover, only variants flagged as “PASSED” by Manta were kept for further interpretation.

Samples submitted by ITHACA, RND, NMD, and GENTURIS were filtered using lists of known disease genes provided by each ERN (3,081, 1,820, 611, 230 genes, respectively, submitted in Laurie et al, 2023). Only SVs affecting at least one exon or located within 5bp of an exon of these genes, were reported to the corresponding submitters for clinical interpretation. Intersection of SVs with short HIGH impact variants (McLaren et al., 2016) and known pathogenic missense variants (Landrum et al., 2020) for identification of compound heterozygous events yielded no additional candidate SVs beyond the disease gene lists. Finally, samples with candidate SVs on more than 5 chromosomes were filtered out, since high numbers of candidate SVs suggested low quality of the DNA or sequencing data rather than the presence of several possibly disease-causing rare SVs on different chromosomes.

Further evaluation of technical quality and potential causality was done individually by the 42 submitting clinicians and research groups. All SV calls affecting genes reported as autosomal dominant (AD) and X-linked (XL, dominant [XLD] or recessive [XLR]) in OMIM (Amberger et al., 2015) with AF less than 4 per cohort (thus, likely affecting members of only one family or few independent patients) were evaluated. Investigation of potential biallelic variants in recessive disease genes, i.e., a combination of an SV with a small heterozygous variant on the other allele, was done by submitters if sufficient expert time was available, which yielded one solved case. Technical validation of calls was undertaken at corresponding facilities. Clear-cut variants were considered as “validation not required”. Such SVs required multiple sources of sequencing signal, such as PE and RD in samples with no obvious quality issues, a match of one affected gene with the disease phenotype and the inheritance patterns, as well as no alternative variants explaining the phenotype.

Visual inspection and quality assessment were done using the IGV genome browser. We generated screenshots of the left and right breakpoints and the complete SV as described in (Demidov et al., 2023). Screenshots accompanying the filtered variants were sent for inspection to the data submitters.

## Results

We have detected, quality-filtered, and annotated SVs in our dataset. SV callsets from unaffected relatives were used for segregation analysis, population-AF based filtering and identification of systematic errors, which helped to dramatically reduce the number of potentially causal SV candidates for in-depth inspection. After automated quality- and annotation-based filtering of the raw SV callsets generated by Manta, 1,404 SV in 868 samples remained in ITHACA, 798 SV in 487 samples in RND, 1,519 SV in 606 samples in NMD and 15 SV in 15 samples in GENTURIS callsets. The distribution of SVs per sample was not uniform, and some samples contained a high number of candidates for clinical interpretation.

Upon expert-inspection, 23 distinct SVs were considered to be causal in 32 out of 6,224 (0.51%) affected individuals, pertaining to 23 unrelated families (Table 1). Eight of these 23 SVs (0.13% of the affected individuals) were not reported by read depth (RD) based CNV detection methods such as ClinCNV (Demidov et al., 2022), ExomeDepth (Plagnol et al., 2012) and Conifer (Krumm et al., 2012) performed in parallel (Demidov et al., 2023) (Table 2). To evaluate the added diagnostic value in comparison with conventional read-depth CNV analysis for ES, we present the identified SVs in three categories.

**Table 1:** The overall number of detected structural variant calls, number of evaluated calls and the diagnostic value increase per ERN.

|  | ERN RND | ERN ITHACA | ERN NMD | ERN GENTURIS |
| --- | --- | --- | --- | --- |
| Number of affected individuals | 2,343 | 1,892 | 1,632 | 357 |
| Number of index patients | 2,200 | 1,821 | 1,499 | 340 |
| Known disease genes in gene list | 1,820 | 3,081 | 611 | 230 |
| Number of candidate variants, after filtering | 798 | 1,404 | 1,519 | 15 |
| Number of samples with SVs, after filtering | 487 | 868 | 606 | 15 |
| Number of solved index patients/all affected patients | 7 (0.32%) / 11 | 9 (0.49%) / 9 | 6 (0.4%) / 9 | 1 (0.29%) / 3 |
| Percentage of causal SVs among investigated SVs | 1.37% | 0.64% | 0.59% | 20% |

**Table 2:** Variants detected via PE or SC signal based SV analysis (Manta) in exomes, considered to be causative for the corresponding rare diseases.

| SIMPLE CNVS |  |  |  |  |  |  |  |  |  |
| --- | --- | --- | --- | --- | --- | --- | --- | --- | --- |
| ERN | Chr | Start | End | Manta Type | IGV evaluation | Gene(s) | Phenostore IDs | Detected by CNV analysis | Length |
| GENTURIS | 16 | 68845794 | 68999240 | BND_0/1 | DEL | CDH1 | P0014615; P0014616; P0014613 | YES | 153446 |
| NMD | 10 | 69917991 | 69950535 | BND_0/1 | TANDEM_DUP (within gene) | MYPN | P0002529 | YES | 32544 |
| ITHACA | 6 | 31630177 | 31657902 | BND_0/1 | DEL | CSNK2B | P0012931 | YES | 27725 |
| NMD | 2 | 179448847 | 179459005 | BND_0/1 | DEL | TTN | P0015420; P0015418 | YES | 10158 |
| RND | 12 | 58014702 | 58024265 | BND_0/1 | DEL | B4GALNT1 | P0015028 | YES | 9563 |
| RND | 16 | 89610020 | 89619318 | BND_1/1 | DEL | SPG7 | P0009735 | YES | 9298 |
| ITHACA | 2 | 162269403 | 162274096 | BND_0/1 | DEL | TBR1 | P0010006 | YES | 4693 |
| ITHACA | 22 | 51160830 | 51163907 | DEL_0/1 | DEL | SHANK3 | P0008274 | NO | 3077 |
| RND | 11 | 66472730 | 66475186 | BND_0/1 | DEL | SPTBN2 | P0011213 | YES | 2456 |
| RND | 3 | 38938611 | 38939384 | BND_0/1 | DEL | SCN11A | P0011371 | NO | 773 |
| NMD | 17 | 48247360 | 48247704 | DEL_1/1 | DEL | SGCA | P0006139; P0006025 | YES | 344 |
| ITHACA | 6 | 1,58E+08 | 1,58E+08 | BND_0/1 | DEL | ARID1B | P0017997 | NO | 245 |
| NMD | 6 | 152485331 | 152485416 | DUP_0/1 | TANDEM_DUP (within gene) | SYNE1 | P0001683 | NO | 85 |
| ITHACA | X | 73749063 | 73749133 | DUP_0/1 | TANDEM_DUP (within gene) | SLC16A2 | P0010004 | NO | 70 |
| ITHACA | 14 | 36987112 | 36987178 | DEL_0/1 | DEL | NKX2-1 | P0007167 | NO | 66 |
| PARTS OF COMPLEX EVENTS |  |  |  |  |  |  |  |  |  |
| RND | 14 | 50878079 | 51132277 | BND_0/1 | INVERSION | ATL1 | P0011754; P0011750; P0011751; P0011753; P0011755 | YES | 254198 |
| RND | X | 153630035 | 153668349 | BND_0/1 | COMPLEX_DUP | ATP6AP1; GDI1; RPL10; TAZ | P0011318 | YES | 38314 |
| ITHACA | 16 | 2214362 | 2229919 | BND_0/1 | DUP (partial) | TRAF7 | P0012480 | YES | 15557 |
| RND | 3 | 11070530 | 11075542 | BND_0/1 | TANDEM_DUP (within gene) plus DEL | SLC6A1 | P0015673 | YES | 5012 |
| NMD | 2 | 179553482 | 179557174 | DUP_0/1 | DEL+DUP | TTN | P0001298; P0001301 | YES | 3692 |
| ITHACA | X | 152958716 | 152959469 | Mix | Retroduplication | SLC6A8 | P0009442 | YES | 753 |
| COPY-NUMBER NEUTRAL |  |  |  |  |  |  |  |  |  |
| ITHACA | 9 | 130887682 | 140727114 | BND_0/1 | INVERSION | EHMT1 | P0019776 | NO | 9839432 |
| NMD | X | 30960717 | 31140070 | BND_1/1 | INVERSION | DMD | P0015754 | NO | 179353 |

First, a detected SV can be classified as a simple deletion or duplication (CNV). 15 distinct SVs in 19 patients were simple CNVs, however, 5 of them were not detected by the CNV detection approaches due to their small size of the affected region (typically, one exon or only part of an exon affected by deletions with overall length ranging from 66 to 3,077 base pairs). The longest CNV, not detected by RD methods, was a 3,077 base-pair long deletion, which only affects 36 base pairs of exon 23 of *SHANK3* (NM_001372044.1), an exon with a total length of 2.2kb. One simple deletion affecting the recessive gene *B4GALNT1* was not reported in RD-based CNV analysis due to an excessive number of CNV candidates in the analysis, but was prioritized in our SV analysis, which produced a much smaller number of calls for expert evaluation. Further inspection of this 9,563bp deletion, removing six exons, revealed that it is detectable with good quality by RD-based CNV callers. Moreover, a missense variant, known to be pathogenic, was detected in the other allele, thus forming a compound heterozygote explaining the phenotype [Laurie at al 2023, submitted]. It is worth mentioning that even for SVs detected in parallel by RD-based CNV callers the accurate information on breakpoint coordinates can facilitate the functional interpretation of the variant, as in e.g. (São José et al., 2023).

The second category comprises variants for which a detected SV can be a part of a complex SV (a combination of deletions, duplications, and inversions), part of which is detectable via standard RD-based CNV analysis. It is rarely possible to detect all breakpoints in this situation using ES data. However, detected SVs in combination with CNVs detected by RD-methods can indicate and potentially better resolve the complex nature of an event. Six variants which were considered causal in 10 patients were complex events, such as a deletion followed by a duplication, or an inversion followed by a duplication.

Finally, a detected SV can be copy-number neutral. We were able to find causal inversions, but no translocations. Two pathogenic inversions were each found in two patients, including one of a size almost 10Mb affecting the *EHMT1* gene (ERN-ITHACA; Fig. 1) and another one of approximately 180kb affecting the *DMD* gene (ERN-NMD). Both inversions occurred in patients with strikingly fitting phenotypes which were explicitly examined for variants in the corresponding genes as candidate genes for several years before being enrolled into the Solve-RD project. The genes are directly affected by inversion breakpoints, hence leading to loss-of-function phenotypes.

**Figure 1:**
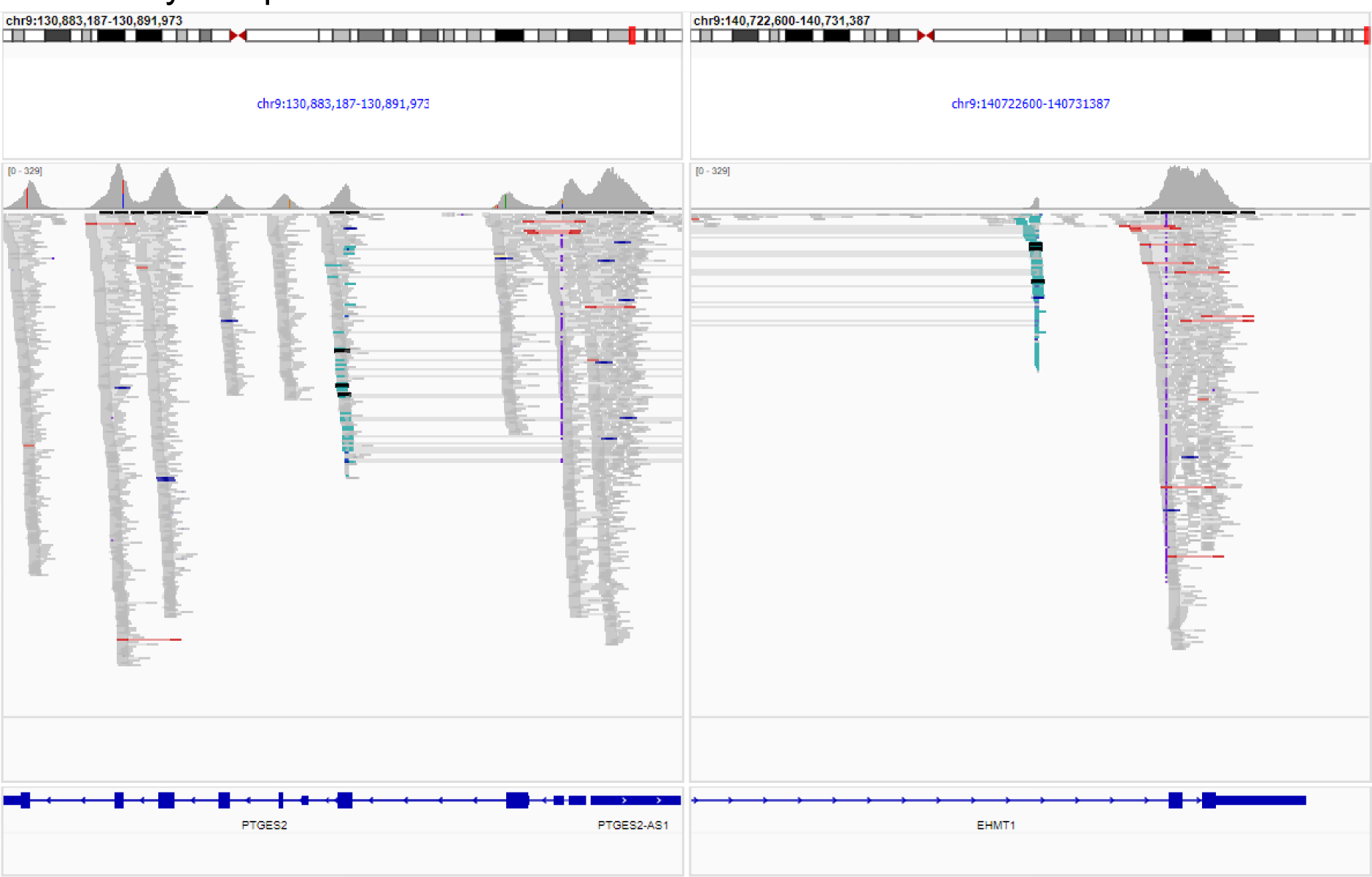
A 9.8Mb-long genomic inversion affecting the penultimate intron in the *EHMT1* gene was detected via structural variants calling in ES data, view in Integrative Genomics Viewer (Robinson et al., 2011).

Thus, the added diagnostic value gained solely from paired-end mapping-based SV detection in the reanalysis of unsolved individuals, was 8 out of 5,825 index patients (0.14%) for pipelines that include a comprehensive read-depth based CNV detection method. This value increases to 0.4% (23/5,825) for pipelines detecting only short variants, hence lacking CNV calls.

## Discussion

Here we demonstrate the utility of PE- and SC-based SV calling in ES data for undiagnosed individuals with a rare disease, using a large cohort with almost 10,000 individuals from the Solve-RD project. Despite the modest overall increase in the diagnostic yield, each successfully diagnosed patient represents a family whose diagnostic odyssey is coming to an end. Especially for patients sequenced years ago, alternative ways for investigating structural variants, such as GS or long-read sequencing, can be unavailable due to financial reasons or lack of DNA samples. Thus, re-analysis approaches using existing ES samples to solve unsolved patients benefit from PE-based SV calling. In a global effort to solve previously unsolved patients, SV calling in ES data could help hundreds of people to find their diagnosis after many years of being undiagnosed.

Even though most of our findings (15 out of 23 unique causal SVs) were also discovered in parallel via read-depth based CNV analysis, SV identification using PE and SC signals should not be considered redundant in such cases. In some patients (6 out of 23 variants) the detected SVs occur next to CNVs detected by RD callers, but knowledge of the SV’s presence is crucial for the interpretation of the rearrangement, uncovering its underlying complexity, and may indicate the necessity for additional molecular analyses such as long read DNA sequencing or RNA sequencing. Furthermore, SV detection in ES, even when it identifies the same variant also found by RD methods, increases the confidence that the variant is a true positive. PE-based SV calling facilitates the validation of CNVs since it provides information about the exact breakpoint coordinates and enables locus-specific PCR amplification and Sanger sequencing. Moreover, we were able to identify eight SVs not identified by RD methods (Demidov et al., 2023), five of which were simple deletions and duplications, two were inversions, and one was a larger deletion forming a compound heterozygote with a SNV in an autosomal recessive disease gene (which was not reported based on RD-calling due to an excessive number of CNV calls in recessive genes).

Notably, both inversions were identified in patients with strikingly matching phenotypes, who underwent multiple rounds of genetic testing of the respective genes before the start of Solve-RD. For example, in one affected individual from an Italian family (GE018), an inversion interrupting the *EHMT1* gene (MIM * 607001) in intron 25 of 26 (g:9:130,887,682-140,727,115, hg19) was detected (see Figure 1). This subject meets the criteria for Kleefstra syndrome type 1 (KS1) (Kleefstra et al., 2009), a well-described syndromic neurodevelopmental condition characterized by psychomotor delay, cognitive impairment, behavioral disorders, facial dysmorphism, abnormal skull shape, abnormalities of hands, and congenital heart defects (Kleefstra et al., 2009; Willemsen et al., 2012). Indeed, this patient showed severe intellectual disability with absence of speech, hand stereotypies, aggressivity, and hypotonia. Additional clinical manifestations consistent with a possible diagnosis of KS1 included dysmorphic features (e.g., sparse eyebrows, short nose, protruding tongue, absence of lateral incisors) and hand abnormalities (syndactyly and drumstick fingers). Considering the suggestive phenotype, genetic testing for KS1 was performed through array comparative genomic hybridization (aCGH) and Sanger sequencing of the *EHMT1* gene, according to the suggestions from the diagnostic guidelines (Kleefstra et al., 2009). However, these tests did not detect any possibly pathogenic variant in the candidate gene, leading to several years of a delay in diagnosing the patient despite the strongly suggestive clinical and phenotypical observations. In this regard, the reanalysis of the genetic data focused on the investigation of structural variants (SV) through the Solve-RD platform was fundamental for the identification of a balanced rearrangement causing the disruption of the *EHMT1* gene, thus providing the pathophysiological link to the genetic condition displayed by the proband of this family. This achievement ended the diagnostic odyssey of this family, finally offering a genetic diagnosis and the chance of targeted clinical management.

In comparison with widely used RD based CNV callers, we cannot estimate the recall rate for detecting variants with PE-based SV calling from ES data, since breakpoints need to affect a targeted region of the exome. For this reason, the chance to successfully identify an SV is independent of its length, as its breakpoints must be covered with a significant number of short reads exhibiting abnormal orientation or insert size. Nonetheless, among our more than 6,000 index patients, five CNVs were detected that were missed by CNV analysis since they were too small. Furthermore, CNV-calling was unable to detect copy-number-neutral events, such as two inversions in our cohort. Moreover, 16 out of 105 unique CNVs found to be causal (Demidov et al., 2023) were also detected in parallel by SV analysis (24 out of 115 solved, candidate or partially explaining the phenotype patients), allowing to better define their breakpoints. Interestingly, this fraction is higher than expected when considering the genomic region covered by ES (<2%), likely because we focus our analysis on SVs and CNVs overlapping known disease genes. Moreover, the chance of detected CNVs to be true positives increases, if they are supported by paired-end and split-read signatures, allowing genetic analysts to evaluate the clinical relevance of the variant without doubts regarding technical quality. Finally, CNVs detected in ES data do not provide breakpoint information that is as accurate as breakpoint locations generated by SV calling, which further simplifies the clinical interpretation of variants. Hence, we conclude that SV calling in ES provides valuable information even for variants already detected with CNV analysis.

A previously published study by (Gardner et al., 2021) in which SVs were analyzed in a cohort of 13,438 probands from the Deciphering Developmental Disorders (DDD) study came to comparable conclusions regarding the fraction of detectable causal SVs. Gardner et al. used a combination of the tool XHMM (Fromer & Purcell, 2014) for RD-based CNV calling and InDelible for split read (SR)-based SV calling, while we combined three RD-based tools with the SV caller Manta. Gardner et al. report 30 unique pathogenic SVs identified by InDelible (0.22%) in addition to 128 CNVs detected by XHMM (0.95%). Interestingly, the fraction of causal CNVs found via the RD-method XHMM is substantially lower compared to the combination of three RD-Methods in Solve-RD (0.95% vs. 1.6%) (Demidov et al., 2023), but in turn the additional diagnostic value of SVs is slightly higher (0.22% vs. 0.14% in our study). Of note, applying InDelible to our cohort (data not shown) did not reveal additional SVs except for 3 mobile element insertions (MEI), which we had already detected using specialized tools(Wijngaard et al., 2023), but missed several causal SVs identified by Manta. We conclude that despite using different patient cohorts and different approaches for the detection of SVs and CNVs, the studies Decipher and Solve-RD produced comparable results regarding the fraction of detectable causal SVs.

As discussed, the PE-based SV calling approach using ES has some weaknesses. We did not perform an evaluation of SV calling sensitivity in ES since the sensitivity is low, as expected by design of targeted sequencing. Usage of Manta or analogous tools for SV detection cannot replace RD-based CNV detection. SV calling should only be considered as an addition, which may increase the diagnostic yield (slightly), but does not guarantee robust detection even of long rearrangements if their breakpoints are intergenic or deep-intronic. Moreover, further improvements and automation of the clinical interpretation of SVs occurring in recessive genes are necessary to reduce the number of reported calls for expert evaluation. This could be achieved by automated phenotypic matching procedures. We plan to apply such an improved procedure during the analysis of the final Solve-RD data freeze and will report the results to the medical genetics community.

In summary, PE and SC based SV calling is a valuable addition to RD-based CNV calling, providing a diagnostics solution to a small but important fraction of rare disease patients.

## Data Availability and Ethics Statement

Data will be deposited at EGA. Accession numbers to be provided. The family (FAM) and participant (P) identifiers used in this manuscript are pseudonymized and known only to the researchers involved In Solve-RD. The Ethics committee of the Eberhard Karl University of Tubingen gave ethical approval for this work.

## Acknowledgement

We want to fully acknowledge the interpretation, evaluation and validation of the candidate structural variants, performed by the following co-authors: Alfons Macaya, Belén Pérez-Dueñas, Miquel Raspall-Chaure, Adam Jackson ((1) Division of Evolution & Genomic Sciences, School of Biological Sciences, Faculty of Biology, Medicine and Health, University of Manchester, Manchester, UK. (2) Manchester Centre for Genomic Medicine, St. Mary’s Hospital, Manchester University NHS Foundation Trust, Health Innovation Manchester, Manchester, UK), Swati Naik (Birmingham Women’s and Children’s Hospital NHS Trust, Birmingham, UK), Giovanni Stevanin (Brain and Spine Institute - ICM, Neurogenetics unit, Sorbonne University, EPHE, Paris, France), Jean-Madeleine de Sainte Agathe (Département de Génétique Médicale, AP-HP.Sorbonne Université, Paris, France), Lukáš Ryba, Markéta Havlovicová (Department of Biology and Medical Genetics, 2nd Faculty of Medicine, Charles University in Prague and Motol University Hospital, Prague, Czech Republic), Rita Horvath (Department of Clinical Neurosciences, University of Cambridge), Michele Pinelli (Department of Molecular Medicine and Medical Biotechnology, University of Naples Federico II, Italy), Nienke J.H. van Os, Bart P.C. van de Warrenburg (Department of Neurology, Radboud university medical center, Nijmegen, The Netherlands), Denommé-Pichon Anne-Sophie, Callier Patrick (FHU TRANSLAD, CHU Dijon, France and GAD team, INSERM UMR1231, Université de Bourgogne-Franche Comté, Dijon, France), Marco Savarese, Mridul Johari (Folkhälsan Research Center, Department of Medical and Clinical Genetics, Medicum, University of Helsinki, Finland), Bruno Dallapiccola, Marco Tartaglia (Genetics and Rare Disease Research Division, Ospedale Pediatrico Bambino Gesù, IRCCS, Rome, Italy), Pauly Martje G. (Institut of Neurogentics, University of Lübeck, Lübeck, Germany), Anna Katharina Sommer (Institute of Human Genetics, Medical Faculty, University of Bonn, Bonn, Germany), Tobias B Haack (Institute of Medical Genetics and Applied Genomics, University of Tübingen, Tübingen, Germany), Ana Topf (John Walton Muscular Dystrophy Research Centre, Translational and Clinical Research Institute, Newcastle University and Newcastle Hospitals NHS Foundation Trust, Newcastle upon Tyne, UK.), Valeria Capra (Medical Genetics Unit, IRCCS Istituto Giannina Gaslini, Genoa, Italy), Lacombe Didier (Medical Genetics, CHU Bordeau; INSERM U1211, Université de Bordeaux), Flavia Pivitera, Chiara Fallerini, Maria Antonietta Mencarelli, Ilaria Longo, Francesca Ariani, Kristina Zguro (Medical Genetics, Med Biotech Hub and Competence Center, Department of Medical Biotechnologies, University of Siena, Italy), Alessandra Renieri (Medical Genetics, Med Biotech Hub and Competence Center, Department of Medical Biotechnologies, University of Siena, Italy, Genetica Medica, Azienda Ospedaliero-Universitaria Senese, Italy), Patrick F. Chinnery (MRC Mitochondrial Biology Unit, Department of Clinical Neurosciences, University of Cambridge), Daniel Natera-de Benito, Andres Nascimento (Neuromuscular Unit, Hospital Sant Joan de Déu, Barcelona, Spain), Aurélien Trimouille (Pathology Department, Bordeaux University Hospital - Inserm U1211-MRGM Univ. Bordeaux), Francina Munell, David Gómez-Andrés, Anna Marcé-Grau (Pediatric Neurology Research Group, Vall d’Hebron Research Institute, Barcelona, Spain), Ben Yaou Rabah (Sorbonne Université, INSERM, Institut de Myologie, Centre de Recherche en Myologie, Database unit, Paris, France), Gisèle Bonne (Sorbonne Université, INSERM, Institut de Myologie, Centre de Recherche en Myologie, Paris, France), Liedewei Van de Vondel (Translational Neurosciences, Faculty of Medicine and Health Sciences, University of Antwerp, Antwerp, Belgium).

We acknowledge bioinformatic analysis done by Burcu Yaldiz (Department of Human Genetics, Radboud Institute for Molecular Life Sciences, Radboud University Medical Centre), Leon Schuetz (Institute of Medical Genetics and Applied Genomics, University of Tübingen, Tübingen, Germany).

## Corporate author list

### Solve-RD consortium

**EKUT:** Olaf Riess^1, 2^, Tobias B. Haack^1^, Holm Graessner^1, 2^, Birte Zurek^1, 2^, Kornelia Ellwanger^1, 2^, Stephan Ossowski^1, 3^, German Demidov^1^, Marc Sturm^1^, Julia M. Schulze-Hentrich^1^, Rebecca Schüle^1, 2^, Jishu Xu^4, 5^, Christoph Kessler^4, 5^, Melanie Kellner^4, 5^, Matthis Synofzik^4, 5^, Carlo Wilke^4, 5^, Andreas Traschütz^4, 5^, Ludger Schöls^4, 5^, Holger Hengel^4, 5^, Holger Lerche^1^, Josua Kegele^6^, Peter Heutink^4, 5^

**RUMC:** Han Brunner^7-9^, Hans Scheffer^7, 8^, Nicoline Hoogerbrugge^7, 10^, Alexander Hoischen^7, 10, 11^, Peter A.C.’t Hoen ^10, 12^, Lisenka E.L.M. Vissers^7, 8^, Christian Gilissen^7, 10^, Wouter Steyaert^7, 10^, Karolis Sablauskas^7^, Richarda M. de Voer^7, 10^, Erik-Jan Kamsteeg^7^, Bart van de Warrenburg^8, 13^, Nienke van Os^8, 13^, Iris te Paske^7,10^, Erik Janssen^7, 10^, Elke de Boer^7, 8^,Marloes Steehouwer^7^, Burcu Yaldiz^7^, Tjitske Kleefstra^7, 8^

**University of Leicester**: Anthony J. Brookes^14^, Colin Veal^14^, Spencer Gibson^14^, Vatsalya Maddi^14^, Mehdi Mehtarizadeh^14^, Umar Riaz^14^, Greg Warren^14^, Farid Yavari Dizjikan^14^, Thomas Shorter^14^

**UNEW:** Ana Töpf^15^, Volker Straub^15^, Chiara Marini Bettolo^15^, Jordi Diaz Manera^15^, Sophie Hambleton^16^, Karin Engelhardt^16^

**MUH:** Jill Clayton-Smith^17, 18^, Siddharth Banka^17, 18^, Elizabeth Alexander^18^, Adam Jackson^17, 18^

**DIJON:** Laurence Faivre^19-23^, Christel Thauvin^19-23^, Antonio Vitobello^21^, Anne-Sophie Denommé-Pichon^21^, Yannis Duffourd^21, 22^, Ange-Line Bruel^21^, Christine Peyron^24, 25^, Aurore Pélissier^24, 25^

**CNAG-CRG:** Sergi Beltran^26, 27^, Ivo Glynne Gut^26, 27^, Steven Laurie^26^, Davide Piscia^26^, Leslie Matalonga^26^, Anastasios Papakonstantinou^26^, Gemma Bullich^26^, Alberto Corvo^26^, Marcos Fernandez-Callejo^26^, Carles Hernández^26^, Daniel Picó^26^, Ida Paramonov^26^, Hanns Lochmüller^26^

**EURORDIS:** Gulcin Gumus^28^, Virginie Bros-Facer^29^

**INSERM-Orphanet:** Ana Rath^30^, Marc Hanauer^30^, David Lagorce^30^,Oscar Hongnat^30^,Maroua Chahdil^30^,Emeline Lebreton^30^

**INSERM-ICM:** Giovanni Stevanin^31-35^, Alexandra Durr^31-34, 36^, Claire-Sophie Davoine^31-35^, Léna Guillot-Noel^31-35^, Anna Heinzmann ^31-34, 37^, Giulia Coarelli^31-34, 37^

**INSERM-CRM:** Gisèle Bonne^38^, Teresinha Evangelista^38^, Valérie Allamand^38^, Isabelle Nelson^38^, Rabah Ben Yaou^38-40^, Corinne Metay^38, 41^, Bruno Eymard^38, 39^, Enzo Cohen^38^, Antonio Atalaia^38^, Tanya Stojkovic^38, 39^

**Univerzita Karlova:** Milan Macek Jr.^42^, Marek Turnovec^42^, Dana Thomasová^42^, Radka Pourová Kremliková^42^, Vera Franková^42^, Markéta Havlovicová^42^, Petra Lišková^43, 44^, Pavla Doležalová^45^

**EMBL-EBI:** Helen Parkinson^46^, Thomas Keane^46^, Mallory Freeberg^46^, Coline Thomas^46^, Dylan Spalding^46^

**Jackson Laboratory**: Peter Robinson^47^, Daniel Danis^47^

**KCL**: Glenn Robert^48^, Alessia Costa^49^, Christine Patch^49, 50^

**UCL-IoN**: Mike Hanna^51^, Henry Houlden^52^, Mary Reilly^51^, Jana Vandrovcova^52^, Stephanie Efthymiou^52^, Heba Morsy^52^, Elisa Cali^52^, Francesca Magrinelli^53^, Sanjay M. Sisodiya^54^, Jonathan Rohrer^55^

**UCL-ICH**, Francesco Muntoni^56, 57^, Irina Zaharieva^56^, Anna Sarkozy^56^

**Universiteit Antwerpen**: Vincent Timmerman^58, 59^, Jonathan Baets^60, 61^, Geert de Vries^59, 60^, Jonathan De Winter^59-61^, Danique Beijer^58-60^, Peter de Jonghe^59, 61^, Liedewei Van de Vondel^58-60^, Willem De Ridder^59-61^, Sarah Weckhuysen^60, 62^

**Uni Naples/Telethon UDP**: Vincenzo Nigro^63, 64^, Margherita Mutarelli^64, 65^, Manuela Morleo^64^, Michele Pinelli^64^, Alessandra Varavallo^64^, Sandro Banfi^63, 64^, Annalaura Torella^63^, Francesco Musacchia^63, 64^, Giulio Piluso^63^

**UNIFE**: Alessandra Ferlini^66^, Rita Selvatici^66^, Francesca Gualandi^66^, Stefania Bigoni^66^, Rachele Rossi^66^, Marcella Neri^66^

**UKB**: Stefan Aretz^67, 68^, Isabel Spier^67, 68^, Anna Katharina Sommer^67^, Sophia Peters^67^

**IPATIMUP**: Carla Oliveira^69-71^, Jose Garcia-Pelaez^69, 70, 72^, Rita Barbosa**-**Matos^69, 70, 73^, Celina São José^69, 70, 72^, Marta Ferreira^69, 70, 74^, Irene Gullo^69-71, 75^, Susana Fernandes^76^, Luzia Garrido^75^, Pedro Ferreira^69, 70, 77^, Fátima Carneiro^69-71, 75^

**UMCG**: Morris A Swertz^78^, Lennart Johansson^78^, Joeri K van der Velde^78^, Gerben van der Vries^78^, Pieter B Neerincx^78^, David Ruvolo^78^, Kristin M Abbott^79^, Wilhemina S Kerstjens Frederikse^79, 80^, Eveline Zonneveld-Huijssoon^79, 81^, Dieuwke Roelofs-Prins^78^, Marielle van Gijn^79, 81^

**Charité**: Sebastian Köhler^82^

**SHU**: Alison Metcalfe^48, 83^

**APHP**: Alain Verloes^84, 85^, Séverine Drunat^84, 85^, Delphine Heron^86, 87^, Cyril Mignot^86, 88^, Boris Keren^86^, Jean-Madeleine de Sainte Agathe^86^

**CHU Bordeaux**: Caroline Rooryck^89^, Didier Lacombe^89^, Aurelien Trimouille^90^

**Spain UDP**: Manuel Posada De la Paz^91^, Eva Bermejo Sánchez^91^, Estrella López Martín^91^, Beatriz Martínez Delgado^91^, F. Javier Alonso García de la Rosa^91^

**Ospedale Pediatrico Bambino Gesù, Rome**: Andrea Ciolfi^92^, Bruno Dallapiccola^92^, Simone Pizzi^92^, Francesca Clementina Radio^92^, Marco Tartaglia^92^

**University of Siena**: Alessandra Renieri^93-95^, Simone Furini^93, 94^, Chiara Fallerini^93, 94^, Elisa Benetti^93, 94^

**Semmelweis University Budapest**: Peter Balicza^96^, Maria Judit Molnar^96^

**University of Ljubljana**, Ales Maver^97^, Borut Peterlin^97^

**University of Lübeck**: Alexander Münchau^98^, Katja Lohmann^99^, Rebecca Herzog^98, 100^, Martje Pauly^98, 99^

**Val d’Hebron Barcelona**: Alfons Macaya^101, 102^, Ana Cazurro-Gutiérrez^101^, Belén Pérez-Dueñas^101^, Francina Munell^101^, Clara Franco Jarava^103, 104^, Laura Batlle Masó^105, 106^, Anna Marcé-Grau^101^, Roger Colobran^103, 104, 107^

**Hospital Sant Joan de Déu Barcelona**: Andrés Nascimento Osorio^108^, Daniel Natera de Benito^108^

**University of Freiburg**: Hanns Lochmüller^109-111^, Rachel Thompson^111^, Kiran Polavarapu^111^, Bodo Grimbacher^112-116^

**University of Oxford**: David Beeson^117^, Judith Cossins^117^

**Folkhälsan Research Centre**: Peter Hackman^118^, Mridul Johari^118^, Marco Savarese^118^, Bjarne Udd^118-120^

**University of Cambridge**: Rita Horvath^121^, Patrick F. Chinnery^121, 122^, Thiloka Ratnaike^123^, Fei Gao^121^, Katherine Schon^121, 124^

**Catalan Institute of Oncology, Barcelona**: Gabriel Capella^125^, Laura Valle^125^

**KU Munich**: Elke Holinski-Feder^126^, Andreas Laner^127^, Verena Steinke-Lange^126^

**TU Dresden**: Evelin Schröck^128^, Andreas Rump^128, 129^

**Koç University:** Ayşe Nazlı Başak^130^

**Ghent University Hospital**: Dimitri Hemelsoet^131, 132^, Bart Dermaut^132-134^, Nika Schuermans^132-134^, Bruce Poppe^132-134^, Hannah Verdin^133^

**University Hospital Meyer, Florence**: Davide Mei^135^, Annalisa Vetro^135^, Simona Balestrini^135, 136^, Renzo Guerrini^135^

**KU Leuven**: Kristl Claeys^137, 138^

**LUMC**: Gijs W.E. Santen^139^, Emilia K. Bijlsma^139^, Mariette J.V. Hoffer^139^, Claudia A.L. Ruivenkamp^139^

**Ludwig Boltzmann Institute for Rare and Undiagnosed Diseases, Vienna:** Kaan Boztug^140-144^, Matthias Haimel^140-142^

**Institute of Pathology and Genetics, Gosselies, Belgium**: Isabelle Maystadt^145, 146^

**Technical University Munich**: Isabell Cordts^147^, Marcus Deschauer^147^

**Neurology/Neurogenetics Laboratory University of Crete, Heraklion, Crete, Greece:** Ioannis Zaganas^148^, Evgenia Kokosali^148^, Mathioudakis Lambros^148^, Athanasios Evangeliou^149^, Martha Spilioti^150^, Elisabeth Kapaki^151^, Mara Bourbouli^151^

**IRCCS G. Gaslini**: Pasquale Striano^152, 153^, Federico Zara^153, 154^, Antonella Riva^153, 154^, Michele Iacomino^154, 155^, Paolo Uva^155^, Marcello Scala^152, 153^, Paolo Scudieri^153, 154^

**Cliniques universitaires Saint-Luc (CUSL)**: Maria-Roberta Cilio^156^, Evelina Carpancea^156^, Chantal Depondt^157^, Damien Lederer^158^, Yves Sznajer^159^, Sarah Duerinckx^160^, Sandrine Mary^158^

**Institute of Human Genetics, University Hospital Essen**: Christel Depienne^161, 162^, Andreas Roos^111, 163, 164^

**University of Luxembourg**: Patrick May^165^

Affiliations

1. Institute of Medical Genetics and Applied Genomics, University of Tübingen, Tübingen, Germany.

2. Centre for Rare Diseases, University of Tübingen, Tübingen, Germany.

3. NGS Competence Center Tübingen (NCCT), University of Tübingen, Tübingen, Germany.

4. Department of Neurodegeneration, Hertie Institute for Clinical Brain Research (HIH), University of Tübingen, Tübingen, Germany.

5. German Center for Neurodegenerative Diseases (DZNE), Tübingen, Germany.

6. Department of Neurology and Epileptology, Hertie Institute for Clinical Brain Research (HIH), University of Tübingen, Tübingen, Germany.

7. Department of Human Genetics, Radboud University Medical Center, Nijmegen, The Netherlands.

8. Donders Institute for Brain, Cognition and Behaviour, Radboud University Medical Center, Nijmegen, The Netherlands.

9. Department of Clinical Genetics, Maastricht University Medical Centre, Maastricht, the Netherlands.

10. Radboud Institute for Molecular Life Sciences, Nijmegen, The Netherlands.

11. Department of Internal Medicine and Radboud Center for Infectious Diseases (RCI), Radboud University Medical Center, Nijmegen, the Netherlands.

12. Center for Molecular and Biomolecular Informatics, Radboud University Medical Center, Nijmegen, the Netherlands.

13. Department of Neurology, Radboud University Medical Center, Nijmegen, The Netherlands.

14. Department of Genetics and Genome Biology, University of Leicester, Leicester, UK.

15. John Walton Muscular Dystrophy Research Centre, Translational and Clinical Research Institute, Newcastle University and Newcastle Hospitals NHS Foundation Trust, Newcastle upon Tyne, UK.

16. Primary Immunodeficiency Group, Translational and Clinical Research Institute, Newcastle University and Newcastle upon Tyne Hospitals NHS Foundation Trust, Newcastle upon Tyne, UK.

17. Division of Evolution, Infection and Genomics, School of Biological Sciences, Faculty of Biology, Medicine and Health, University of Manchester, Manchester M13 9WL, UK.

18. Manchester Centre for Genomic Medicine, St Mary’s Hospital, Manchester University Hospitals NHS Foundation Trust, Health Innovation Manchester, Manchester M13 9WL, UK.

19. Dijon University Hospital, Genetics Department, Dijon, France.

20. Dijon University Hospital, Centre of Reference for Rare Diseases: Development disorders and malformation syndromes, Dijon, France.

21. Inserm - University of Burgundy-Franche Comté, UMR1231 GAD, Dijon, France.

22. Dijon University Hospital, FHU-TRANSLAD, Dijon, France.

23. Dijon University Hospital, GIMI institute, Dijon, France.

24. University of Burgundy-Franche Comté, Dijon Economics Laboratory, Dijon, France.

25. University of Burgundy-Franche Comté, FHU-TRANSLAD, Dijon, France.

26. CNAG-CRG, Centre for Genomic Regulation (CRG), The Barcelona Institute of Science and Technology, Baldiri Reixac 4, Barcelona 08028, Spain.

27. Universitat Pompeu Fabra (UPF), Barcelona, Spain.

28. EURORDIS-Rare Diseases Europe, Sant Antoni Maria Claret 167 - 08025 Barcelona, Spain.

29. EURORDIS-Rare Diseases Europe, Plateforme Maladies Rares, 75014 Paris, France.

30. INSERM, US14 - Orphanet, Plateforme Maladies Rares, 75014 Paris, France.

31. Institut National de la Santé et de la Recherche Medicale (INSERM) U1127, Paris, France.

32. Centre National de la Recherche Scientifique, Unité Mixte de Recherche (UMR) 7225, Paris, France.

33. Unité Mixte de Recherche en Santé 1127, Université Pierre et Marie Curie (Paris 06), Sorbonne Universités, Paris, France.

34. Institut du Cerveau - ICM, Paris, France.

35. Ecole Pratique des Hautes Etudes, Paris Sciences et Lettres Research University, Paris, France.

36. Centre de Référence de Neurogénétique, Hôpital de la Pitié-Salpêtrière, Assistance Publique-Hôpitaux de Paris (AP-HP), Paris, France.

37. Hôpital de la Pitié-Salpêtrière, Assistance Publique-Hôpitaux de Paris (AP-HP), Paris, France.

38. Sorbonne Université, Inserm, Institut de Myologie, Centre de Recherche en Myologie, F-75013 Paris, France.

39. AP-HP, Centre de Référence de Pathologie Neuromusculaire Nord, Est, Ile-de-France, Institut de Myologie, G.H. Pitié-Salpêtrière, F-75013 Paris, France.

40. Institut de Myologie, Equipe Bases de données, G.H. Pitié-Salpêtrière, F-75013 Paris, France.

41. AP-HP, Unité Fonctionnelle de Cardiogénétique et Myogénétique Moléculaire et Cellulaire, G.H. Pitié-Salpêtrière, F-75013 Paris, France.

42. Department of Biology and Medical Genetics, Charles University Prague-2nd Faculty of Medicine and University Hospital Motol, Prague, Czech Republic.

43. Department of Paediatrics and Inherited Metabolic Disorders, First Faculty of Medicine, Charles University and General University Hospital in Prague, Prague, Czech Republic.

44. Department of Ophthalmology, First Faculty of Medicine, Charles University and General University Hospital in Prague, Prague, Czech Republic.

45. Centre for Paediatric Rheumatology and Autoinflammatory Diseases, Department of Paediatrics and Inherited Metabolic Disorders, 1st Faculty of Medicine, Charles University and General University Hospital in Prague, Czech Republic.

46. European Bioinformatics Institute, European Molecular Biology Laboratory, Wellcome Genome Campus, Hinxton, Cambridge, United Kingdom.

47. Jackson Laboratory for Genomic Medicine, Farmington, CT 06032, USA.

48. Florence Nightingale Faculty of Nursing, Midwifery & Palliative Care, King’s College, London, UK.

49. Society and Ethics Research, Connecting Science, Wellcome Genome Campus, Hinxton, UK.

50. Genomics England, Queen Mary University of London, Dawson Hall, EC1M 6BQ, London, UK.

51. MRC Centre for Neuromuscular Diseases and National Hospital for Neurology and Neurosurgery, UCL Queen Square Institute of Neurology, London, UK.

52. Department of Neuromuscular Diseases, UCL Queen Square Institute of Neurology, London, UK.

53. Department of Clinical and Movement Neurosciences, UCL Queen Square Institute of Neurology, University College London, WC1N 3BG.

54. Department of Clinical and Experimental Epilepsy, UCL Queen Square Institute of Neurology, London, UK.

55. Dementia Research Centre, Department of Neurodegenerative Disease, UCL Queen Square Institute of Neurology, London, UK.

56. Dubowitz Neuromuscular Centre, UCL Great Ormond Street Hospital, London, UK.

57. NIHR Great Ormond Street Hospital Biomedical Research Centre, London, United Kingdom.

58. Peripheral Neuropathy Research Group, University of Antwerp, Antwerp, Belgium.

59. Laboratory of Neuromuscular Pathology, Institute Born-Bunge, University of Antwerp, Antwerpen, Belgium.

60. Translational Neurosciences, Faculty of Medicine and Health Sciences, University of Antwerp, Belgium.

61. Neuromuscular Reference Centre, Department of Neurology, Antwerp University Hospital, Antwerpen, Belgium.

62. VIB-CMN, Applied and Translational Neurogenomics Group.

63. Dipartimento di Medicina di Precisione, Università degli Studi della Campania “Luigi Vanvitelli”, Napoli, Italy.

64. Telethon Institute of Genetics and Medicine, Pozzuoli, Italy.

65. Istituto di Scienze Applicate e Sistemi Intelligenti “E.Caianiello” - ISASI -CNR.

66. Unit of Medical Genetics, Department of Medical Sciences, University of Ferrara, Italy.

67. Institute of Human Genetics, Medical Faculty, University of Bonn, Bonn, Germany.

68. Center for Hereditary Tumor Syndromes, University Hospital Bonn, Bonn, Germany.

69. i3S - Instituto de Investigação e Inovação em Saúde, Universidade do Porto, Portugal.

70. IPATIMUP - Institute of Molecular Pathology and Immunology of the University of Porto, Portugal.

71. Faculty of Medicine, University of Porto, Portugal.

72. Doctoral Programme in Biomedicine, Faculty of Medicine, University of Porto, Portugal.

73. Doctoral Programme in BiotechHealth, School of Medicine and Biomedical Sciences, University of Porto, Portugal.

74. Doctoral Programme in Computer Science, Faculty of Sciences, University of Porto, Portugal.

75. CHUSJ, Centro Hospitalar e Universitário de São João, Porto, Portugal.

76. Departament of Genetics, Faculty of Medicine, University of Porto, Portugal.

77. Faculty of Sciences, University of Porto, Portugal.

78. Department of Genetics, Genomics Coordination Center, University Medical Center Groningen, University of Groningen, Groningen, The Netherlands.

79. Department of Genetics, University Medical Center Groningen, University of Groningen, Groningen, The Netherlands.

80. ERN-GENTURIS.

81. ERN-RITA: European Reference Network for Immunodeficiency, Autoinflammatory, Autimmune and Paediatric Rheumatic diseases, Utrecht, Netherlands.

82. Ada Health GmbH, Karl-Liebknecht-Str. 1, 10178 Berlin, Germany.

83. College of Health, Well-being and Life-Sciences, Sheffield Hallam University, Sheffield, UK.

84. Dept of Genetics, Assistance Publique-Hôpitaux de Paris - Université de Paris, Robert DEBRE University Hospital, 48 bd SERURIER, Paris, France.

85. INSERM UMR 1141 “NeuroDiderot”, Hôpital Robert DEBRE, Paris, France.

86. Department of Genetics, Assistance Publique-Hôpitaux de Paris - Sorbonne Université, Pitié-Salpêtrière University Hospital, 83 Boulevard de l’Hôpital, Paris, France.

87. Reference center of rare diseases “intellectuel disability of rare causes”, Paris, France.

88. Institut du Cerveau (ICM), UMR S 1127, Inserm U1127, CNRS UMR 7225, Sorbonne Université, 75013, Paris, France.

89. Univ. Bordeaux, MRGM INSERM U1211, CHU de Bordeaux, Service de Génétique Médicale, F-33000 Bordeaux, France.

90. Laboratoire de Génétique Moléculaire, Service de Génétique Médicale, CHU Bordeaux – Hôpital Pellegrin, Place Amélie Raba Léon, 33076 Bordeaux Cedex, France.

91. Institute of Rare Diseases Research, Spanish Undiagnosed Rare Diseases Cases Program (SpainUDP) & Undiagnosed Diseases Network International (UDNI), Instituto de Salud Carlos III, Madrid, Spain.

92. Molecular Genetics and Functional Genomics, Ospedale Pediatrico Bambino Gesù, IRCCS, Rome, Italy.

93. Med Biotech Hub and Competence Center, Department of Medical Biotechnologies, University of Siena, Italy.

94. Medical Genetics, University of Siena, Italy.

95. Genetica Medica, Azienda Ospedaliero-Universitaria Senese, Italy.

96. Institute of Genomic Medicine and Rare Diseases, Semmelweis University, Budapest, Hungary.

97. Clinical Institute of Genomic Medicine, University Medical Centre Ljubljana, Slovenia.

98. Institute of Systems Motor Science, University of Lübeck, Ratzeburger Allee 160, 23562, Lübeck, Germany.

99. Institute of Neurogenetics, University of Lübeck, Ratzeburger Allee 160, 23562, Lübeck, Germany.

100. Department of Neurology, University Hospital Schleswig Holstein, Ratzeburger Allee 160, 23562, Lübeck, Germany.

101. Pediatric Neurology Research Group, Vall d’Hebron Research Institute, Universitat Autònoma de Barcelona, Barcelona, Spain.

102. Institute of Neuroscience, Universitat Autònoma de Barcelona, Barcelona, Spain.

103. Diagnostic Immunology Research Group, Vall d’Hebron Research Institute (VHIR), Barcelona, Spain.

104. Immunology Division, Genetics Department. Vall d’Hebron University Hospital (HUVH), Barcelona, Spain.

105. Infection in Immunocompromised Pediatric Patients Research Group, Vall d’Hebron Research Institute (VHIR), Barcelona, Spain.

106. Pediatric Infectious Diseases and Immunodeficiencies Unit, Vall d’Hebron University Hospital (HUVH),Barcelona, Spain.

107. Immunology Unit. Department of Cell Biology, Physiology and Immunology. Autonomous University of Barcelona (UAB), Bellaterra, Spain.

108. Neuromuscular Disorders Unit, Department of Pediatric Neurology. Hospital Sant Joan de Déu, Barcelona, Spain

109. Department of Neuropediatrics and Muscle Disorders, Medical Center, Faculty of Medicine, University of Freiburg, Freiburg, Germany.

110. Centro Nacional de Análisis Genómico (CNAG-CRG), Center for Genomic Regulation, Barcelona Institute of Science and Technology (BIST), Barcelona, Spain.

111. Children’s Hospital of Eastern Ontario Research Institute, University of Ottawa, Ottawa, Canada.

112. Institute for Immunodeficiency, Center for Chronic Immunodeficiency (CCI), Medical Center, Faculty of Medicine, Albert-Ludwigs-University of Freiburg, Germany.

113. Clinic of Rheumatology and Clinical Immunology, Center for Chronic Immunodeficiency (CCI), Medical Center, Faculty of Medicine, Albert-Ludwigs-University of Freiburg, Germany.

114. DZIF – German Center for Infection Research, Satellite Center Freiburg, Germany.

115. CIBSS – Centre for Integrative Biological Signalling Studies, Albert-Ludwigs University, Freiburg, Germany.

116. RESIST – Cluster of Excellence 2155 to Hanover Medical School, Satellite Center Freiburg, Germany.

117. Nuffield Department of Clinical Neurosciences, University of Oxford, UK.

118. Folkhälsan Research Centre and Medicum, University of Helsinki, Helsinki, Finland.

119. Tampere Neuromuscular Center, Tampere, Finland.

120. Vasa Central Hospital, Vaasa, Finland.

121. Department of Clinical Neurosciences, University of Cambridge, Cambridge, UK.

122. Medical Research Council Mitochondrial Biology Unit, University of Cambridge, Cambridge, UK.

123. Department of Paediatrics, University of Cambridge, Cambridge, UK.

124. East Anglian Medical Genetics Service, Cambridge University Hospitals NHS Foundation Trust, Cambridge, UK.

125. Bellvitge Biomedical Research Institute (IDIBELL), Barcelona, Spain.

126. Medizinische Klinik und Poliklinik IV – Campus Innenstadt, Klinikum der Universität München, Munich, Germany.

127. MGZ - Medical Genetics Center, Munich, Germany.

128. Institute of Clinical Genetics, University Hospital Carl Gustav Carus, Technical University Dresden, Dresden, Germany.

129. Center for Personalized Oncology, University Hospital Carl Gustav Carus, Technical University Dresden, Dresden, Germany.

130. Koç Universıty,School of Medicine, Translational Medicine Research Center, KUTTAM-NDAL Istanbul Turkey.

131. Dpt. of Neurology, Ghent University Hospital, Ghent, Belgium.

132. Program for Undiagnosed Rare Diseases (UD-PrOZA), Ghent University Hospital, Ghent, Belgium.

133. Center for Medical Genetics, Ghent University Hospital, Ghent, Belgium.

134. Department of Biomolecular Medicine, Faculty of Medicine and Health Sciences, Ghent University, Ghent, Belgium.

135. Neuroscience Department, Children’s Hospital A. Meyer-University of Florence, 50139, Florence, Italy.

136. Department of Clinical and Experimental Epilepsy, UCL Queen Square Institute of Neurology, and Chalfont Centre for Epilepsy, Gerrard Cross, UK.

137. Department of Neurology, University Hospitals Leuven, Leuven, Belgium.

138. Laboratory for Muscle Diseases and Neuropathies, Department of Neurosciences, and Leuven Brain Institute (LBI), KU Leuven - University of Leuven, Leuven, Belgium.

139. Department of Clinical Genetics, Leiden University Medical Center, Leiden, The Netherlands.

140. Ludwig Boltzmann Institute for Rare and Undiagnosed Diseases, Vienna, Austria.

141. St. Anna Children’s Cancer Research Institute (CCRI), Vienna, Austria.

142. CeMM Research Center for Molecular Medicine of the Austrian Academy of Sciences, Vienna, Austria.

143. Department of Pediatrics and Adolescent Medicine, Medical University of Vienna, Vienna, Austria.

144. St. Anna Children’s Hospital, Department of Pediatrics and Adolescent Medicine, Medical University of Vienna, Vienna, Austria.

145. Centre de Génétique Humaine, Institut de Pathologie et de Génétique, Gosselies, Belgium.

146. Département de Médecine, Université de namur (Unamur), Namur, Belgique.

147. Department of Neurology, Klinikum rechts der Isar, Technical University Munich, Munich, Germany.

148. Neurology / Neurogenetics Laboratory University of Crete, Heraklion, Crete, Greece.

149. Aristotle University of Thessaloniki, Thessaloniki, Greece.

150. 1st Department of Neurology, Aristotle University of Thessaloniki, University General Hospital of Thessaloniki, AHEPA, Thessaloniki, Greece.

151. Neurochemistry and Biomarker Unit, 1st Department of Neurology, School of Medicine, National and Kapodistrian University of Athens, Eginition Hospital, Athens, Greece.

152. Pediatric Neurology and Muscular Disease Unit, IRCCS Istituto Giannina Gaslini, Genoa, Italy.

153. Department of Neurosciences, Rehabilitation, Ophthalmology, Genetics, Maternal and Child Health, University of Genoa, Genoa, Italy.

154. Unit of Medical Genetics, IRCCS Istituto Giannina Gaslini, Genoa, Italy.

155. Clinical Bioinformatics, IRCCS Istituto Giannina Gaslini, Genoa, Italy.

156. Pediatric Neurology Department, Saint-Luc University Hospital, Université Catholique de Louvain, Brussels, Belgium.

157. Neurology Department, Erasme Hospital, Université Libre de Bruxelles, Bruxelles, Belgium.

158. Institute of Pathology and Genetics, Charleroi, Belgium.

159. Human Genetics Department, Saint-Luc University Hospital, Université Catholique de Louvain, Brussels, Belgium.

160. Institute of Interdisciplinary Research in Human and Molecular Biology, Human Genetics, IRIBHM, Université Libre de Bruxelles, Brussels, Belgium.

161. Institute of Human Genetics, University Hospital Essen, University Duisburg-Essen, Essen, Germany.

162. Institut du Cerveau et de la Moelle épinière (ICM), Sorbonne Université, UMR S 1127, Inserm U1127, CNRS UMR 7225, F-75013 Paris, France.

163. Department of Pediatric Neurology, Developmental Neurology and Social Pediatrics, Children’s Hospital University of Essen, Essen, Germany.

164. Department of Neurology, Heimer Institute for Muscle Research, University Hospital Bergmannsheil, Ruhr-University Bochum, 44789 Bochum, Germany.

165. Luxembourg Centre for Systems Biomedicine, University of Luxembourg, Esch-sur-Alzette, Luxembourg.

